# Emergent effects of contact tracing robustly stabilize outbreaks

**DOI:** 10.1101/2021.02.25.21252445

**Authors:** Seyfullah Enes Kotil

## Abstract

Covid-19 neither dissolved nor got out of control over a year. In many instances, the new daily cases exhibit an equilibrium at a meagre percentage of the population. Seemingly impossible due to the precise cancellation of positive and negative effects. Here, I propose models on real-world networks that capture the mysterious dynamics. I investigate the contact-tracing and related effects as possible causes. I differentiate the impact of contact-tracing into three—one direct and two emergent—effects: isolation of the documented patient’s direct infectees (descendants), isolation of non-descendant infectees, and temporary isolation of susceptible contacts. Contrary to expectation, isolation of descendants cannot stabilize an equilibrium; based on current data, the effect of the latter two are necessary and greater in effect overall. The reliance on emergent effects shows that even if contact-tracing is 100% efficient, its effect on the epidemic dynamics would be dependent. Moreover, This newly characterized dynamic claims that all outbreaks will eventually show such stable dynamics.

## Introduction

Modelling the observed COVID-19 dynamics is vital for understanding the inner-workings of an epidemic. If current models cannot recreate observable phenomena, it is an indication that different factors are at play. We see at least two modes of COVID-19 dynamics from public data. In countries like China, cases increase and then decrease because of harsh lockdowns. Subsequently, the epidemic ceases to exist[1–3, 6]. In other countries like Germany, Italy, and Poland, the epidemic continues to live with stable daily positives for a considerable long time, Fig 1a. The latter countries tend to be more relying on contact tracing strategy. Also, they have extensive isolation and quarantine procedures. The cause of the “equilibrium state” is unknown and unmodeled. Naturally, the way to battle with it is also unclear.

**Fig. 1.**
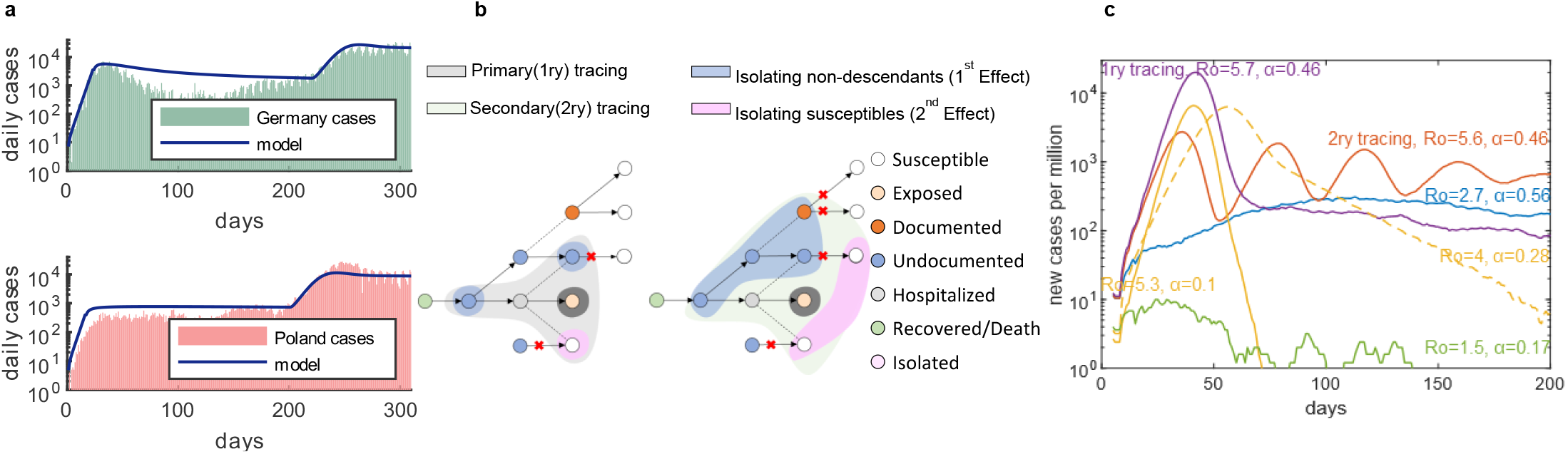
Four distinct spontaneous dynamics can be observed for COVID-19. **a**, In Germany and Poland and many more countries, the covid-19 outbreak is showing stable new daily cases. The ODE model can fit the data, equations (3), (Methods). **b**, Spread of an epidemic depicted like a branching process, where the arrows indicate infection while dotted lines indicate contacts. Two indirect effect of contact-tracing—isolation of non-descendants (blue shade) and isolation of susceptible contacts (pink shade)—halts other branches (infections). The primary contact tracing (grey shade) halts fewer infections than secondary tracing (green shade). **c**, There are four distinct dynamics: stable (blue), oscillatory stable (orange), stability reached after a large wave (purple), eradicated (green), growth and decay (solid yellow). Although we see unstable phase abundantly, the emergent effects attenuate growth and elongate decay phase in almost all simulations (dotted yellow).

SIR (Susceptible-Infected-Resistance) models and other models have been used to understand COVID-19 dynamics [4,5,7–9]. Substantial progress was made predicting the basic reproduction number (*Ro*) [10], and strategies to flatten the curve [11–15]. However, SIR and related models do not have a stable equilibrium: even true for models with contact-tracing. The epidemic can still only exponentially grow or decay, equation (1–2). Moreover, branching process models with “forward tracing “[16]—tracing of descendants—show no difference in equilibrium dynamics. In all of the formulations mentioned above, the birth and death of the branches (infections) are independent. In reality, the branches are not independent: an isolated contact can halt another branch, Fig 1b. In another view, the death rate of other branches increases as the number of branches are discovered, while the birth rate does not—eventually leading to a stable equilibrium.

I investigate two indirect effects that can induce such stability: 1) direct removal of non-descendant COVID-19 positives among the contacts of a documented patient, 2) Temporary isolation of susceptible contacts which prevents possible non-descendants, Fig 1b. Both effects serve the same purpose: decreasing the infectivity of a non-descendants, by removal (the first effect), or by prevention (the second effect). Two effects are naturally occurring. Here, I identify this stable phase as robustly occurring for the natural course of an epidemic.

## Results

This paper investigates the equilibrium state first by an updated system of ODEs, then by agent-based models with random networks and real-world networks. All simulation has populations compartmentalized into five parts; susceptible (S), exposed (E), undocumented (U), documented (D), Hospitalized (H), Recovered/Death (RD), (Supplementary Fig. 1). I assume that the undocumented and documented compartments are infectious.

### The updated ODE model exhibits robust emergence of stability

I first investigate the equilibrium state by an updated system of ordinary differential equations (ODEs), equations (3): the stability is possible, and the model can successfully fit the data, Fig 1a. However, The required traced contacts per documented patient are high to reanimate the dynamics in the data, parameters section, Supplementary Fig. 2, Fig. 1a. The ODE is a well-mixed system. But, the two indirect effects depend on the interrelations between the players. Contrary to ODE modelling, a documented individual’s contacts are not equivalent to a random sample among the population. Individuals are interwoven with the fabric of society. Nevertheless, analysis of ODE results shows three significant findings:

**Fig. 2.**
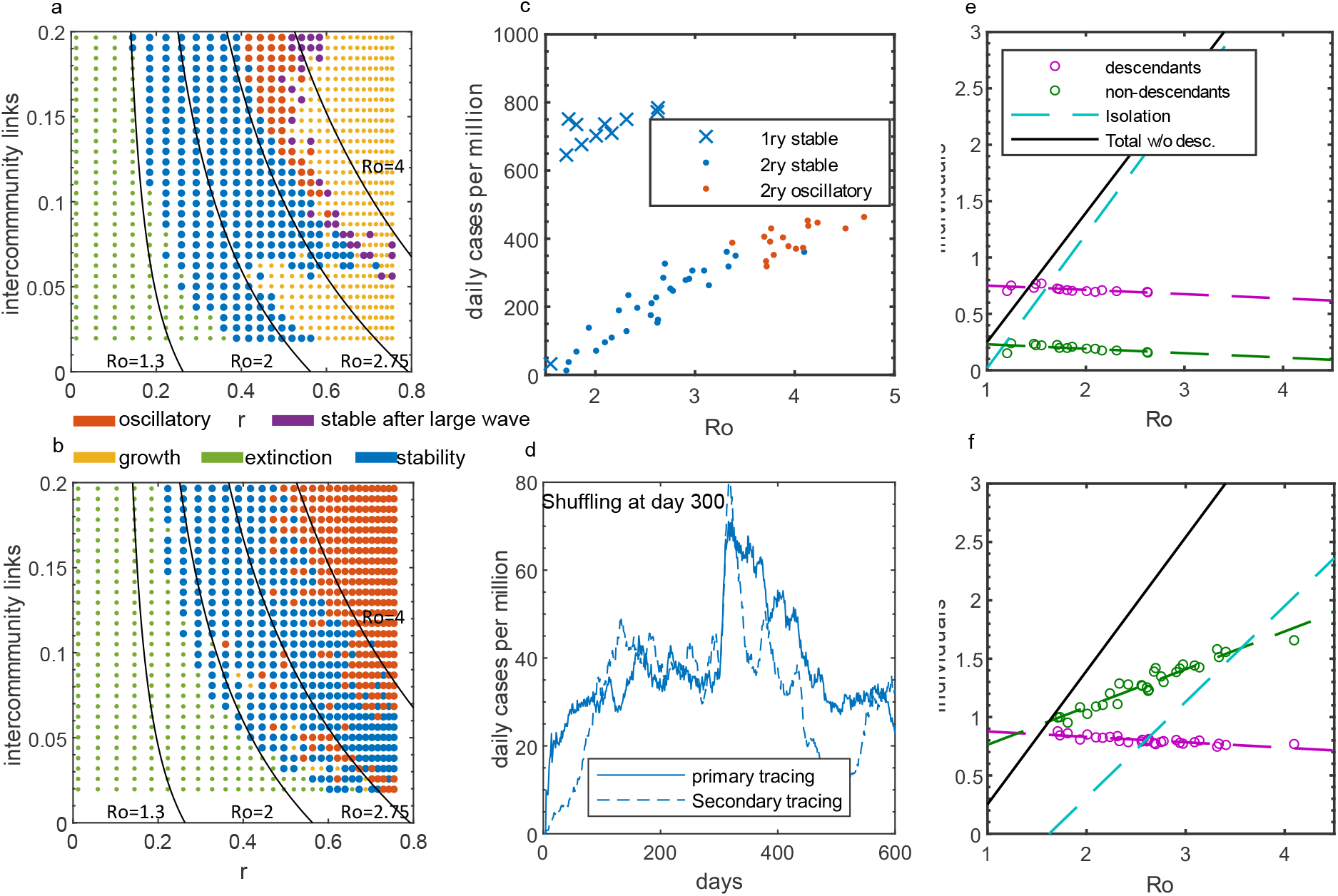
isolation of susceptible contacts is the dominant force that induces stability. **a**, Dependence of four phases on hourly transmission rate (r) and percentage of inter-community links (primary tracing). **b**, dependence for secondary tracing. **c**, The number of daily cases increases linearly with Ro for primary and secondary tracing. There is a 6-fold difference when primary and secondary tracing is compared, Supplementary Fig. 9b. **d**, Population-wide mobilization leads to a wave of infection. The population was shuffled at day 300 for one day. **c**, Classification of the three forces of contact tracing. Isolation of susceptible individuals predominates in simulations with primary tracing when Ro is higher than 2. **d**, removal of non-descendant individuals predominate at intermediate Ro for secondary tracing.

1. The number of total non-descendants that should be removed, or prevented follows a very simple equation, (Ro-1)/(α·T), where Ro is the basic reproduction number, α is the documentation ratio. T is tracing efficiency, Supplementary Fig. 3, equation (4).
2. The number of daily cases at the stability is inversely proportional to contacts isolated per documented, k, equation (5).
3. The existence of the equilibrium state increases with lower Ro, Supplementary Fig. 2.

Even if an outbreak begins with a high Ro (basic reproduction number), the effective reproduction number will decrease and leads the dynamics towards a stable regime due to humans’ natural reaction.

**Fig. 3.**
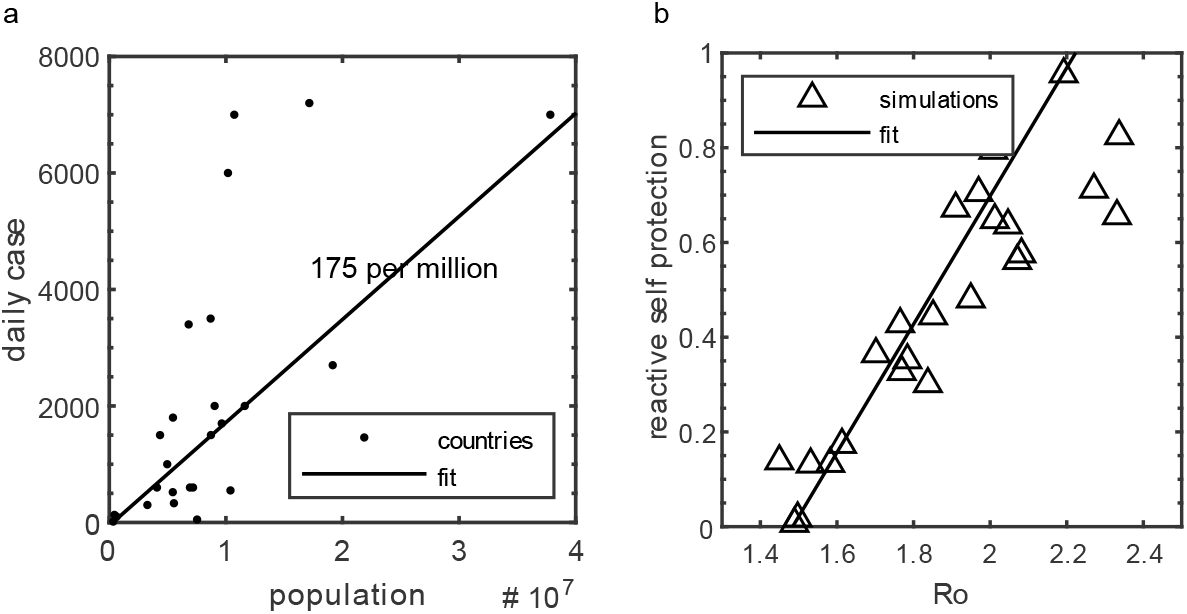
Reactive self-protection is a stabilizing factor that can give rise to observed data. **a**, Daily cases for 25 countries versus their population size. A fit indicates there are 175 cases per million on average. **b**, Simulations with higher values for basic reproduction numbers require high reactive self-protection. The protective behaviour—decreasing transmission rate —is engaged when contact is discovered.

Furthermore, the isolated proportion (by the second effect) of the population equals the herd immunity level, *1-1/Ro*, when the first effect (direct removal of non-descendants) is disabled. In contrast, when both effects are enabled, every removed non-descendant decreases the isolated percentage, equation (6). At the stability, the first effect is directly measurable by counting. However, the second effect can be inferred by comparing the needed total, equation (4), versus the computed, or observed, non-descendants, equation (7).

### An agent-based model simulated on random networks shows four distinct dynamics

Thousands of randomly generated networks were simulated. The random networks show four different phases: eradication, stable, stable with oscillations, stable after large infection wave, Fig. 1d. The latter three dynamics are emergent: it only occurs when players are interdependent. As expected from equation (4), the most predictive parameters were the documentation ratio, *α*, and the basic reproduction number, *Ro*, Supplementary Fig. 4. In addition to *α* and *Ro*, the stable phase depends on clustering, the abundance of inter and intracommunity links, variability in link frequency in a non-trivial way, Supplementary Fig. 5 and 6. Another critical parameter is the tracing radius, the number of traced contacts per individuals. In agent-based modelling, the contacts per individual is not a tuneable parameter. Hence, a high number of traced contacts per individual is achieved by simulating secondary tracing, tracing all direct (primary) contacts’ contacts. For subsequent simulations, I use a point estimation of 0.45 for the documentation ratio [17]. For this limit, primary tracing shows stability at lower *Ro*; meanwhile, the secondary tracing sustains stability, or oscillatory stability, for a wide range of *Ro*.

**Fig. 4.**
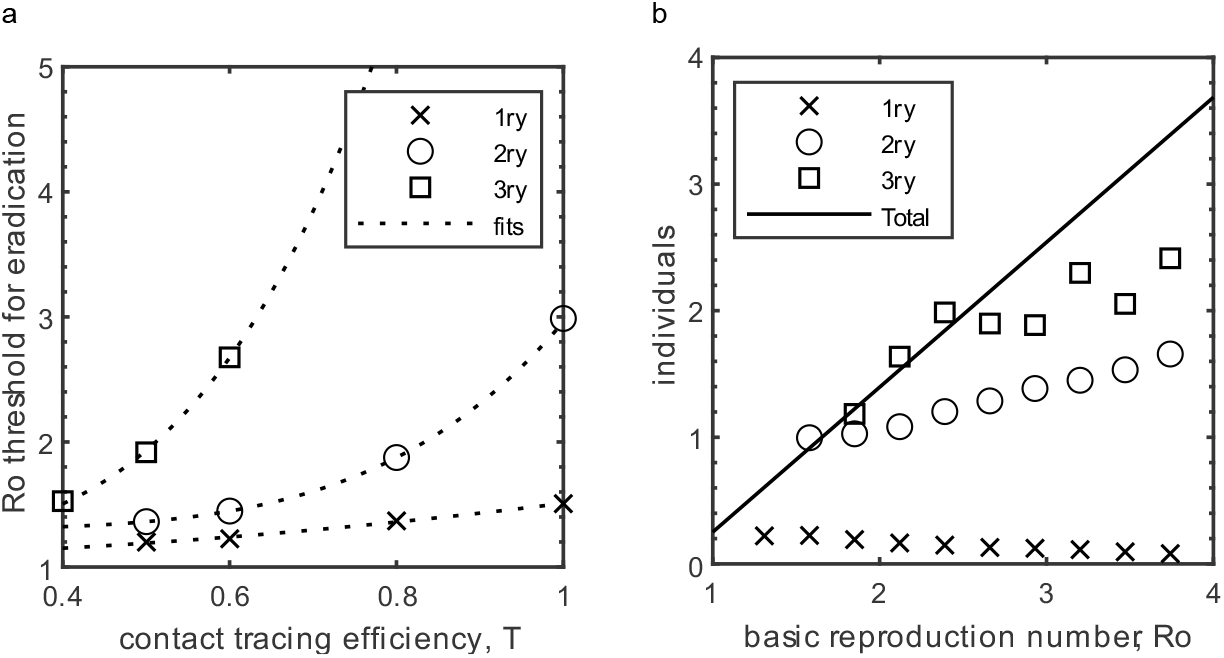
Eradication thresholds increase the most when secondary tracing is performed instead of primary tracing. **a**, Eradication threshold for increasing contact tracing efficiency and the tracing radius. **b**, Isolating non-descendants more than the required total for stability leads to eradication. A major increase in isolation of non-descendants is observed when secondary tracing is implemented.

### Real-world networks show similar emergent behavior

Next, I simulate the real-world network not to force a particular topology. The real-world network contains only 459 people. Unfortunately, the stabilities at very low percentages cannot be observed. Then, I reconstructed a network of 900000 people by replicating the real network, where each replica is analogous to a community, Supplementary Fig. 7 and Methods. Simulations are defined by two independent parameters: transmission probability per hour (*r*), and percentage of inter-community links. Both parameters influence the effective reproduction number. The r decreases with social distancing, mask use and similar precautions, while the intercommunity links decrease by not meeting with people. As consistent with ODE and random networks, an epidemic must experience the equilibrium state, before it can be eradicated. Therefore, the equilibrium state is experienced robustly and spontaneously for both primary and secondary tracing Fig 2a and b.

Like observations from the ODEs, the equilibrium state-level increases linearly with *Ro*, Fig 2c, Supplementary Fig. 8 and 9. For the same parameters, secondary tracing exhibits 6-fold lower equilibrium point than the simulations with the primary tracing, Supplementary Fig. 10a. The isolated percentage for secondary tracing is much higher than the corresponding level for primary tracing, Supplementary Fig. 10b. Expectedly, the conditions for stability are less stringent for real networks compared to other models.

### Network topology influences emergent local behavior

A documented patient is likely to be closer to other COVID-19 positives. Contacts of a patient are not equivalent to a random sample. I test this hypothesis by randomly sampling the same number of contacts compared to the number of non-descendant contacts. The descendants are isolated as before. Strikingly, the isolation levels increase, Supplementary Fig. 11. This phenomenon indicates the local structure of the network is very important. The local structure could be disturbed by effects like mobility. Major acts of shuffling in holidays can induce surges, Fig 2d, Supplementary Fig. 12, 13.

### The isolation of non-descendants is the dominant force compared to removal of descendants and removal of susceptibles

The number of grandchildren that must be isolated for stability was directly counted, Supplementary Fig. 14, (methods). At the stability, the relative importance of the three effects (removal of descendants and the two indirect effects) of contact tracing varies with Ro and the tracing radius. Strikingly, the removal of descendants is comparable to the other forces. Unfortunately, the contact-tracing procedures are designed for tracing descendants. Moreover, for simulations with *Ro* between 2 and 3, temporary isolation of susceptible and the removal of non-descendants are the dominant force for primary and secondary tracing, respectively, Fig 2e, f.

### The primary tracing can be insufficient for reanimating data, and the secondary tracing is not practiced

Many countries had a stability level of around 175 cases per million in winter of 2021[1], Fig 3a. The *Ro* that is corresponding to that is around 1.8, for the primary tracing. However, the robustness of stability is not comparable. Moreover, the inferred Ro would be very close to extinction. Therefore, the primary tracing, alone, cannot explain the data. Although the secondary tracing is not practised, it also does not enable flairs of infection at the considered parameter range. Probably, the real society lies somewhere between the primary and the secondary tracing.

### Reactive self-protective behaviour and Cluster management could be a major contributing factor

The insufficiency of primary tracing leads to consideration of other factors that are at play. I consider human behaviour as a contributing factor. People are well connected and could get information about COVID-19 status of their acquaintances. The information spreads through the network. Possibly, the information could lead them to engage in self-protective behaviour, temporarily lowering their transmission probability. I modelled this mode of action as lowering their original transmission rate proportionally: the primary contacts are isolated as usual, but the secondary contacts receive a transmission penalty. This effect is a stabilizing effect, Supplementary Fig. 15a. The levels seen in Fig. 3a can be reached with various levels of reactive protective behaviour while Ro is between 1.6 and 2.4, Fig 3b.

Next, I consider cluster management as a possible effect. Here, the cluster management is modelled by isolating a whole community (459 individuals) after a predefined number of people are discovered. The cluster management can eradicate a disease if performed very effectively. However, cluster management has minimal effect on the daily cases in secondary tracing: the low frequency of cases identifies clusters difficult, Supplementary Fig 15d. Strikingly, cluster management does not induce stability, Supplementary Fig. 15c. For cluster management to induce stability, the discovery of one cluster should increase another cluster’s discoverability, which may happen. Countries like Japan [18] and South Korea are very proficient in cluster management; they have an abnormally low number of stable daily cases and had days when there were no cases.

### An equilibrium state is attainable even without contact tracing

The protective behaviour can be extended to primary contacts. In doing so, a stable state can be reached. Here, the protective behaviour is applied to both primary and secondary contacts: self-protective behaviour of 60% can induce stability at Ro equals 2.4, Supplementary Fig. 15b.

### Eradication of the equilibrium state

Changes in Ro cannot unstabilize the stable condition, rather the number of daily cases changes. Decreasing Ro decreases the daily cases continuously and eventually crosses a threshold, Ro, eradicating the disease. The Ro threshold increases with higher orders of tracing, Fig 4a. Overall criteria are that the removed non-descendant individuals per documented patient must exceed the amount required for stability. The most prominent jump in removing non-descendants occurs at secondary tracing: mostly attributable to ancestor’s other descendants’ isolation, Fig 4b. Although unrealistic, tertiary tracing eradicated disease at very high Ro. However, the increase is mostly attributable to excessive isolation.

Eradication by vaccination is the current hope of humanity. The criteria for vaccination is to bring the Ro below Ro for eradication.

## Discussion

This work identifies a new emergent dynamic of epidemics. The existence of a stable or oscillatory equilibrium characterizes the dynamic. Strikingly, the new dynamic turns an outbreak into a chronic problem: it cannot overtake the population or be eradicated. Firth et al. earlier showed stability for COVID-19 to require secondary tracing, with most of the population isolated [19]. Another study showed the stability to occur at very low Ro [20]. Here, I show that the natural course of the epidemic robustly experiences stability (or oscillatory stability), even for primary tracing, which exhibits a low percentage of the population as isolated. Furthermore, the stability can even be achieved without contact-tracing: other effects like reactive protective behaviour can induce stability. Fundamentally, I show the isolation or prevention of non-descendants is the causal forces: making the stability an emergent phenomenon.

The used real network is enlarged artificially. The change of size is performed in the most non-disturbing way possible. Nevertheless, more complete networks could yield more accurate results. The existence of the stability is not sensitive to the network topology: although highly influenced. The direct comparison to real data suggests that the primary tracing cannot be enough—other effects like reactive behaviour due to concerning COVID-19 experiences of contacts. The real data is successfully fitted. However, further work is needed to single out a contributor.

The identification of the emergent forces implies that the effect of contact-tracing is more complicated than expected. At the stability, the indirect effects are at least as impactful compared to the isolation of descendants. Thus, even if 100% efficiency for contact tracing is obtained by complete adaptation of tracker apps—the adaptation is far less [21]—the effect on the dynamics can be dominated by other factors. For instance, mobility was shown to correlates with the disease dynamics [22,23]. Here, mobility’s effect on the dynamics is also exerted through the two emergent forces. Unfortunately, the policies are ignoring the two forces that are mentioned in this work.

In this equilibrium, the disease has an effective reproduction number higher than one, but it is kept at bay by the contact-tracing or other related effects. The existence of equilibrium ensures that the performed social distancing is insufficient. Reopening and vaccination strategies are intense areas of research [24,26]. Here, I also provided a threshold to eradicate the disease. The eradication in this paper is more indicative of a disruption of the stable state. In reality, eradication is a more complex problem.

The new emergent equilibrium dynamics is applicable to the social phenomenon as well. Any modality with an appeal of following but is somehow feared or frowned upon can robustly converge to an equilibrium.

## Methods

### ODE models

Standard SEIR-like model for COVID-19 dynamics:

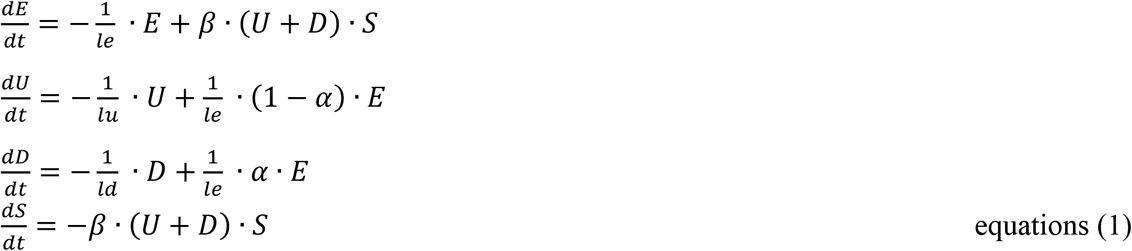

Where *E, U, D* and *S* are the percentage of the population that is Exposed, Undocumented, Documented, Susceptible, respectively. Moreover, *le, ld*, and *lu* are the time interval for being in exposed, documented, and undocumented state, respectively. Also, the *β* is the daily transmission rate, and *α* is the documentation ratio.

Modelling contact tracing with SIR models requires that for each documented patient, its descendants are subtracted from their respective equations. Phenomenologically, contact tracing reduces the infectivity of the documented COVID-19 positives. A simple introduction could be factored into the *r, r* · (1 − *T*) where T is the level of contact tracing. An utterly successful contact tracing is akin to dynamics where only undocumented positives transmit the disease. Standard SEIR-like model with “forward contact tracing” for COVID-19 dynamics:

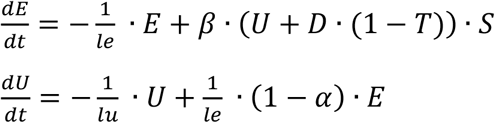

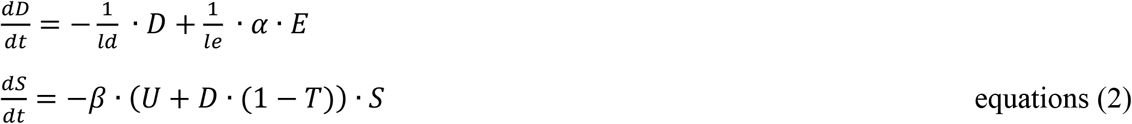

Where *T* is the contact tracing success ratio. The equilibrium conditions for models with and without contact tracing, show no difference in equilibrium dynamics. The epidemic can still only exponentially grow or decay. The equilibrium state is still unstable. The contact tracing modelled in this form cannot explain the stability that we see from the data.

The two effects are modelled with the terms containing *e1* and *e2* for effect 1 and 2, respectively. The updated ODE model is given in equations (3):

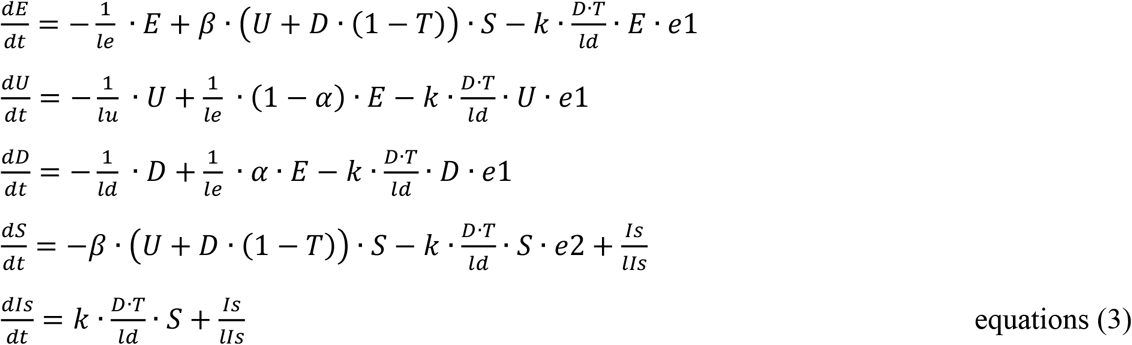

Where *e1* and *e2* are the effectiveness of the two indirect effects, removal of non-descendants, isolation of susceptibles, respectively. k is the number of individuals isolated per successfully traced and documented patient. *lIs* is the day limit for staying in quarantine, and the Is is the isolated percentage.

To quantify the requirements for the stability, I set the e2 to 0. The total non-descendants that need to be removed can be counted directly. To simplify equations, the removal of undocumented and documented individuals are omitted. The majority of the COVID-19 positives are the exposed individuals.

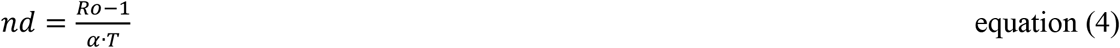

The above simplifications were used to calculate the equilibrium level.

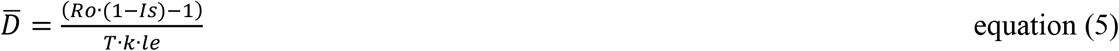

Where the 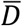 is the average daily documented individuals.

The isolated percentage decreases with number of removed non-descendants, derived from the same formulations of equation (5).

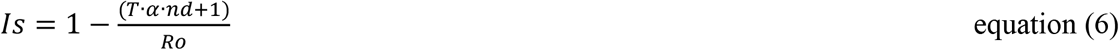

The effect of the isolation of susceptibles is hard to gauge directly. It is an implicit question: what is the probability of infection for an isolated susceptible individual, if that person was not isolated? Nevertheless, the effect 2 can be gauged by subtracting the observed non-descendant removal from the total that is required.

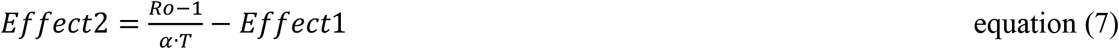

### Construction and simulation of random networks

Total of 17000 trajectories was simulated. Every simulation contains 250000 agents. The agents are linked randomly. Each simulation is defined by the number of links per agent and transmission probability. All trajectories are simulated for 150 days: 2-hour intervals simulate each day. At each time point, the contacts are chosen randomly. Basic reproduction number is estimated directly by counting the descendants of a discovered agent.

### Classification of trajectories into the four phases

The trajectories are divided into four distinct clusters: stable, oscillatory stable, stability reached after a large infection wave, eradicated, growth and decay. The classification is made automatically. First, trajectories that did not grow are marked as eradication. Then, the logarithmic rate change for the last 50 days is measured. The threshold for stability is given as the doubling time, or half-life (0.02) must be more than 34.5 days. If the trajectory is stable, then it is checked for oscillations. Trajectories with 30% deviation from the mean are flagged as oscillatory. If a trajectory is stable or oscillatory, then it is checked if there was a large flair. When the total infected is less than (1-1/Ro)0.8, then it is flagged as, stability with large flair. The remaining trajectories are flagged as growth trajectories. Although not flagged as stable, these trajectories show less growth and elongated decay time compared to trajectories that lack contact tracing.

### Construction and simulation of real-world network

The Haslemere dataset was generated by previous work [5]. The data tracks 1272 individuals. 468 of 1272 individuals had data. Here only 321 individuals are used: ones that have at least a half-hour of contacts in 3 days. The data includes pairwise connections every 5 minutes. This network has two limitations: the total number of individuals is small, a small percentage of the whole population is tracked. Tracking of a small percentage of individuals leads to a fragmented network. To alleviate fragmentation, I take all connections less than 5 meters. The size of the network is solved by replicating the network. Each replica is analogous to a community. Also, Intercommunity links were chosen to make communities behave as parallel universes. The links preserve the wiring by linking the same respective contacts across communities, Supplementary Fig. 7. Intercommunity links are chosen at random. The simulations are made hourly. At each hour, the corresponding 5-minute connections are executed with a constant transmission probability. There three days in the Haslemere data: Thursday, Friday and Saturday. The total simulations take 201 days, where three days are repeated. This repetition induces a little predictable noise. The trajectories are smoothed by moving average for further analysis.

### Quantifying the strength of the three forces of Contact tracing

Isolation of contacts can be divided into three categories: isolating descendants, isolating non-descendants and isolating susceptibles. Descendants are the individuals who are infected by the discovered patient. Non-descendants are the individuals who are infected by others but isolated because of another patient. The number of isolated descendants and non-descendants per discovered patient can be directly counted. However, the effect of isolation of the susceptibles is an implicit question: What is the probability of an isolated susceptible individual to be infected, if it was not isolated?. Nevertheless, it can be inferred by subtracting the counted non-descendants from the total that is required. The total non-descendants that should be prevented or isolated can also be directly counted. Trajectories where no susceptibles are isolated, the counted isolated non-descendants, at the stability, are equivalent to the total that is required. The numerically computed “total” is fitted by a linear line, Supplementary Fig. 8. Thus, the counted non-descendants, from normal trajectories, are subtracted from the fitted function to calculate the effect of isolating susceptibles.

### Randomly isolating individuals

The number of contacts that should be isolated is calculated for every time point, then the same number of randomly chosen contacts are isolated in addition to isolating all randomly. The descendants and non-descendants are isolated, but the susceptible contacts are not isolated. Instead, the same number of susceptibles are isolated from the population randomly. **Mobility**. The identities of all agents are shuffled at day 200 for a complete day (16-time intervals). Meanwhile, all necessary tracing is carried out.

#### Reactive self-protective behavior

Individuals that have contacts that are infected engage in behaviour that effectively reduces their transmission rate. The behaviour is referred to as reactive self-protective behaviour. Reduction of transmission rate decreases the probability to be infected, and the probability of infecting others. The outcome of contacts that both engage in self-protective behaviour is modelled as a product. Primary contacts are isolated, but the secondary contacts engage in self-protective behaviour. The contacts, as before, have a 20 per cent chance of not being found.

Simulations without any contact tracing, but has self-protective behaviour for primary and secondary contacts, can still show stability. The implementation is similar, primary contacts are not isolated, but all contacts, primary and secondary, engage in reactive self-protective behaviour.

#### Estimating stable daily cases from the data

Data from 25 countries were used to estimate the daily cases at stability. The countries were Poland, Romania, Netherlands, Belgium, Czechia, Greece, Portugal, Hungary, Austria Serbia, Switzerland, Hong Kong, Paraguay, Bulgaria, Lebanon, Finland, Slovakia, Norway, Ireland, Panama, Croatia, Bosnia Herzegovina, Luxembourg, Malta, and Iceland. The number of cases was taken from the data in late fall of 2020 and winter of 2021. The data was taken manually.

#### Estimating the extend of reactive self-protective behaviour according to data

Total of 180 combinations of transmission rate and the number of inter-community links were selected. For each setting, 6 levels of [0, 0.2, 0.4, 0.6, 0.8, 1] reactive self-protective behavior was simulated. The reactive self-protective behaviour that brings the simulation to the observed daily cases (176 per million) was determined. Pairs of Ro (influenced by transmission rate and intercommunity links) and self-protective behaviour was plotted, a linear line was fitted for illustration.

### Cluster management

Communities are monitored for documented patients. If a threshold of documented patients is observed within the last 14 days, the whole community (321) is isolated. In this setting, identification of clusters does not induce the discovery of others. Other implementations can easily be performed.

### Eradication

Simulations with different tracing efficiency and tracing radius (primary, secondary, tertiary) were performed. The highest Ro that eradicated the disease is determined. It is important to note that higher-order tracing increases the tracing efficiency of primary contacts. Even if a primary contact was missed, that same contact could be traced as secondary or tertiary. When the level of effective primary tracing is considered, the eradication Ro is not very different at modest effective primary tracing, Supplementary Fig. 16.

### Parameters

**General:** *le*=3.5 days, *lu*=3 days, *ld*=5days. *α*=0.45, *e1*=1, *e2*=1.

**Fig. 1a**. Germany: *k*=180 per documented, *βsummer*=0.51 per day, *D1*=220^th^ day, *βfall*=1 per day; Poland *k*=200 per documented, *βsummer*=0.51 per day, *D1*=200^th^ day, *βfall*=1 per day.

**Fig. 1c**. *Tracing efficiency (T)=*0.8.

**Fig. 2a**. *T*=0.65.

**Fig. 2d. Primary:** *r*=0.29, **Secondary:** *r*=0.39.

**Fig. 2e and f**. inter-community links 10%, *T*=0.65

**Fig. 3b** inter-community links [0.16, 0.13, 0.16, 0.10, 0.13, 0.16, 0.7, 0.10, 0.13, 0.16, 0.07, 0.10, 0.13, 0.16, 0.04, 0.13, 0.10, 0.16, 0.07, 0.04, 0.13, 0.10, 0.07, 0.04, 0.10, 0.07, 0.04, 0.07, 0.04, 0.04] paired with *r* [0.28, 0.30, 0.31, 0.32, 0.33, 0.35, 0.36, 0.36, 0.37, 0.38, 0.41, 0.41, 0.41, 0.41, 0.42, 0.44, 0.45, 0.45, 0.46, 0.48, 0.48, 0.49, 0.51, 0.53, 0.53, 0.55, 0.58, 0.60, 0.63, 0.67]

**Fig. 4a** inter-community links 10%

**Fig. 4b** inter-community links 10%, *T*=0.65

**Supplementary Fig 3**. *e1*=1, *e2*=0

**Supplementary Fig 10**. inter-community links 10%, *T*=0.65

**Supplementary Fig 11a**. inter-community links 10%, *T*=0.65, *r*= 0.89

**Supplementary Fig 11b**. inter-community links 10%, *T*=0.65

**Supplementary Fig 14**. inter-community links 10%, *T*=0.65

**Supplementary Fig 15**. *T*=0.65, inter-community links 10%, *r*= 0.57

**Supplementary Fig 16**. inter-community links 10%

## Data Availability

Available on request

## Reporting Summary

Further information on research design is available in the Nature Research Reporting Summary linked to this article.

## Code availability

The MATLAB scripts used will be made available

## Data availability

The datasets generated during this study will be amde available

## Acknowledgements

This work was supported by TUBITAK, 2232 - International Fellowship for Outstanding Researchers, Project number 11C244. All the results are in sole responsibility of the author. I thank H. Tunc, Z. Sari, B. Darendeli, R. Nashebi and I. Erol for insightful discussions

## Author contributions

S.E.K. designed the study, analysed the data, performed the simulations, and wrote the manuscript.

