## Supplementary material for "Emergent effects of contact tracing robustly stabilize outbreaks"

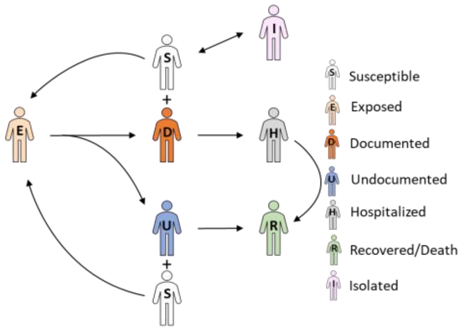


**Supplementary figure 1| Schematic of the disease states.** Schematic of the disease states

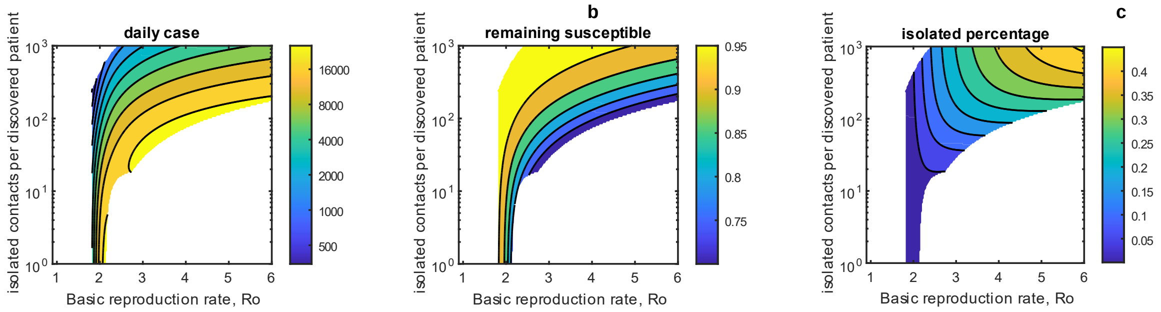


**Supplementary figure 2| Phase planes for the ODE model. a**, Phase plane with contours for the daily cases. **b**, Phase plane with contours for the remaining susceptible population by 200 days. **c**, Phase plane with contours for isolated percentage at the stability.

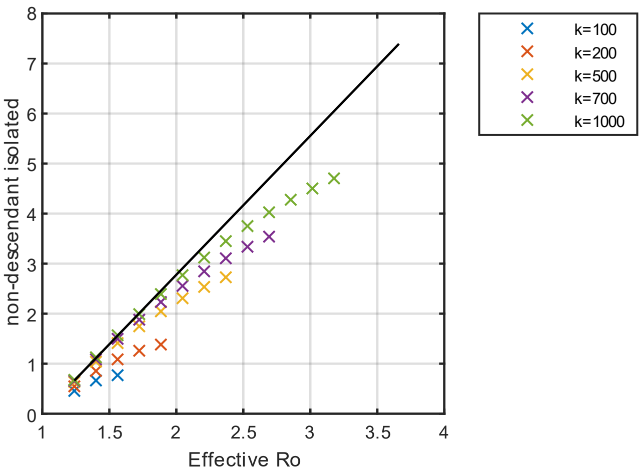


**Supplementary figure 3| Total Non-Grandchildren that must be removed for stability. a**, Number of non-descendant that should be removed for stability increases linearly with Ro (basic reproduction number). The dependence is (Ro-1)/(α T).

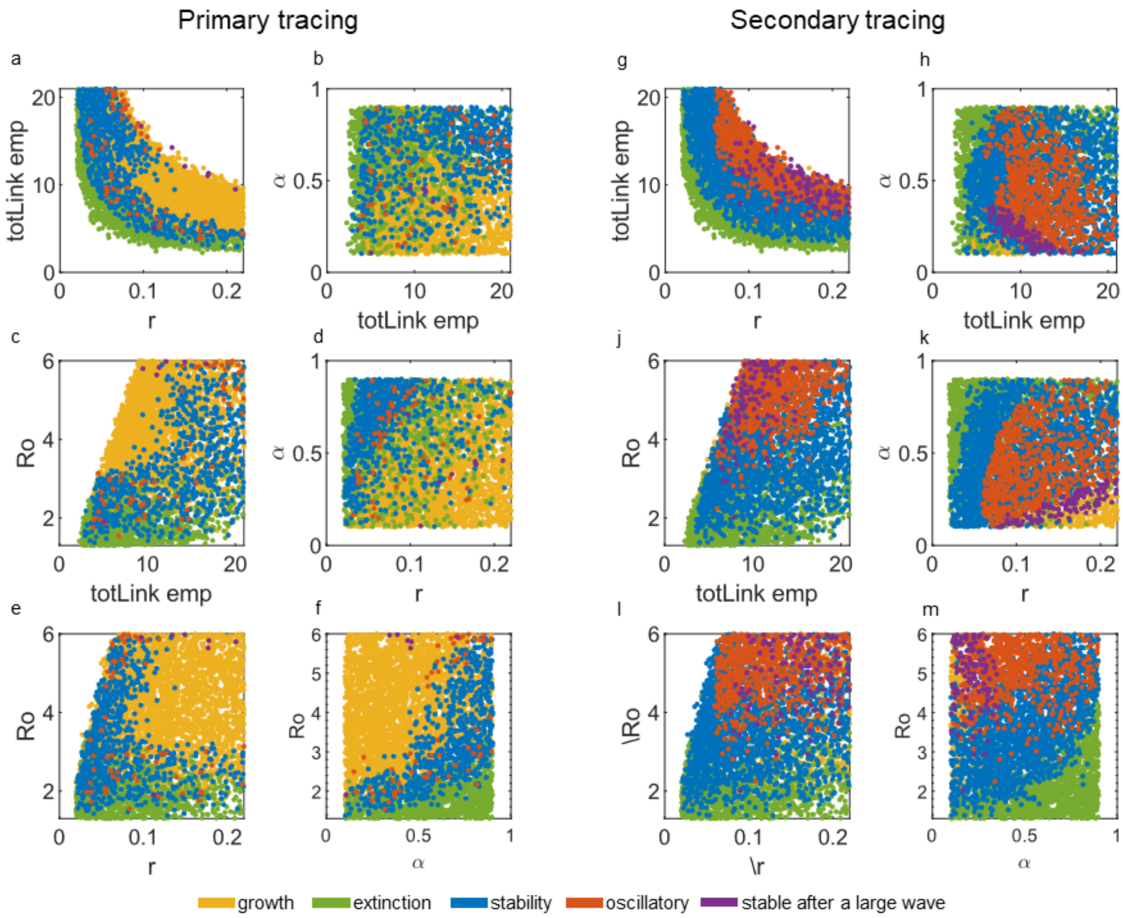


**Supplementary figure 4| Phase planes for the agent-based model with random networks.** A total of 5000 simulations for 250000 people were made. Subfigures from a to f are associated with primary tracing, while the subfigures from g to m are associated with secondary tracing. **a**, Dependence of four phases on the hourly transmission rate and number of links. Links are randomly assigned. The "number of links" refers to the number of connections each agent makes. b, number of links vs documentation ratio. **c**, number of links vs Ro. d, Transmission rate vs documentation ratio. **e**, Transmission rate vs Ro. f, Documentation ratio vs Ro. g, **h**, **j**, **k**, **l**, **m** same dependencies as a to f.

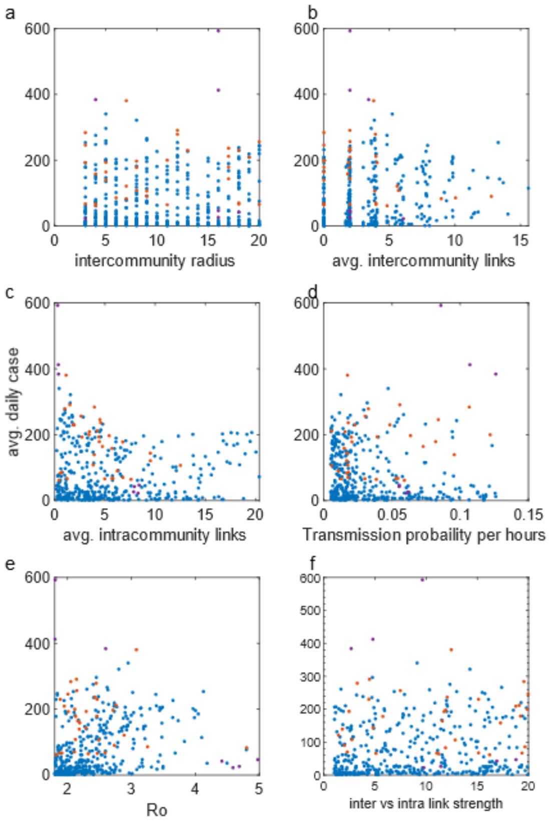


**Supplementary figure 5| Dependence of daily cases on network topology.** All subfigures have simulations with primary tracing. The documentation ratio is 0.45. Total of 5000 trajectories with 250000 agents was performed. Some correlation is observed with the different parameters. However, the dependence is not one dimensional. The most predictive parameter is Ro. **a**, Dependence of daily cases on radius of the community. Two types of links are present in these simulations, inter and intracommunity links. Firstly, the population is divided into non-overlapping subgroups as communities. The members are the square of the "radius of the community". The links inside the communities are the Intralinks, where the links between communities are the interlinks. The interlinks are chosen at random. **b**, Dependence of daily cases on Intralinks. **c**, Dependence of daily cases on interlinks. **d**, Dependence of daily cases on transmission probability. **e**, Dependence of daily cases on Ro. f, Dependence of daily cases on the strength of interlinks vs Intralinks.

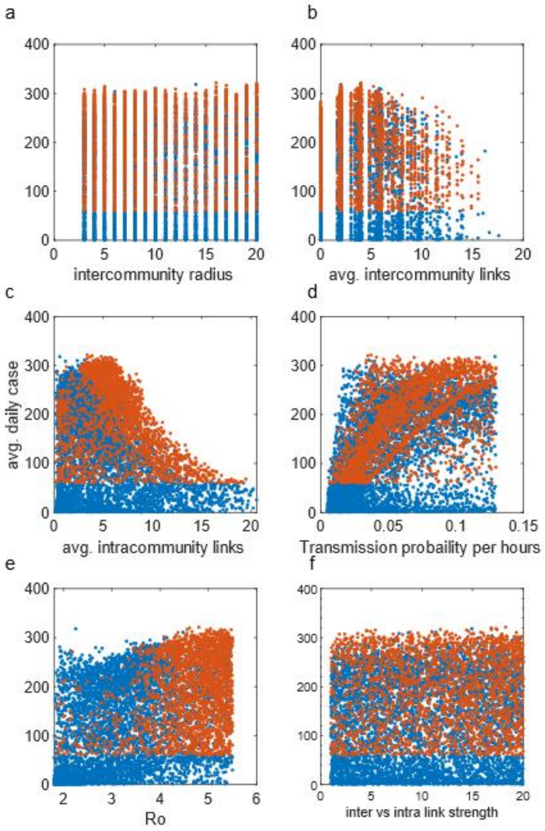


**Supplementary figure 6| Dependence of daily cases on network topology.** All subfigures have simulations with secondary tracing. The documentation ratio is 0.45. Total of 10000 trajectories with 250000 agents was performed. Some correlation is observed with the different parameters. However, the dependence is not one dimensional. The most predictive parameter is the transmission probability. **a**, Dependence of daily cases on the radius of the community. Two types of links are present in these simulations, inter and intracommunity links. Firstly, the population is divided into non-overlapping subgroups as communities. The members are the square of the "radius of the community". The links inside the communities are the Intralinks, where the links between communities are the interlinks. The interlinks are chosen at random. **b**, Dependence of daily cases on Intralinks. **c**, Dependence of daily cases on interlinks. **d**, Dependence of daily cases on transmission probability. **e**, Dependence of daily cases on Ro. f, Dependence of daily cases on the strength of interlinks vs Intralinks.

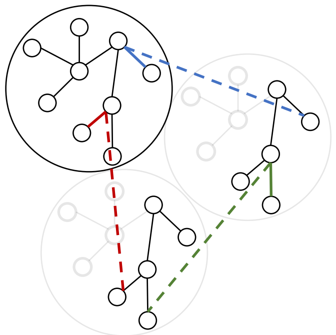

**Supplementary figure 7| The enlarged network consists of parallel universes(communities).** , Schematic of the enlarged real-network. To preserve the topology, the intercommunity links (dashed) are between the corresponding ones in the original pairs (solid coloured) (see methods for more information).

Supplementary figure 8

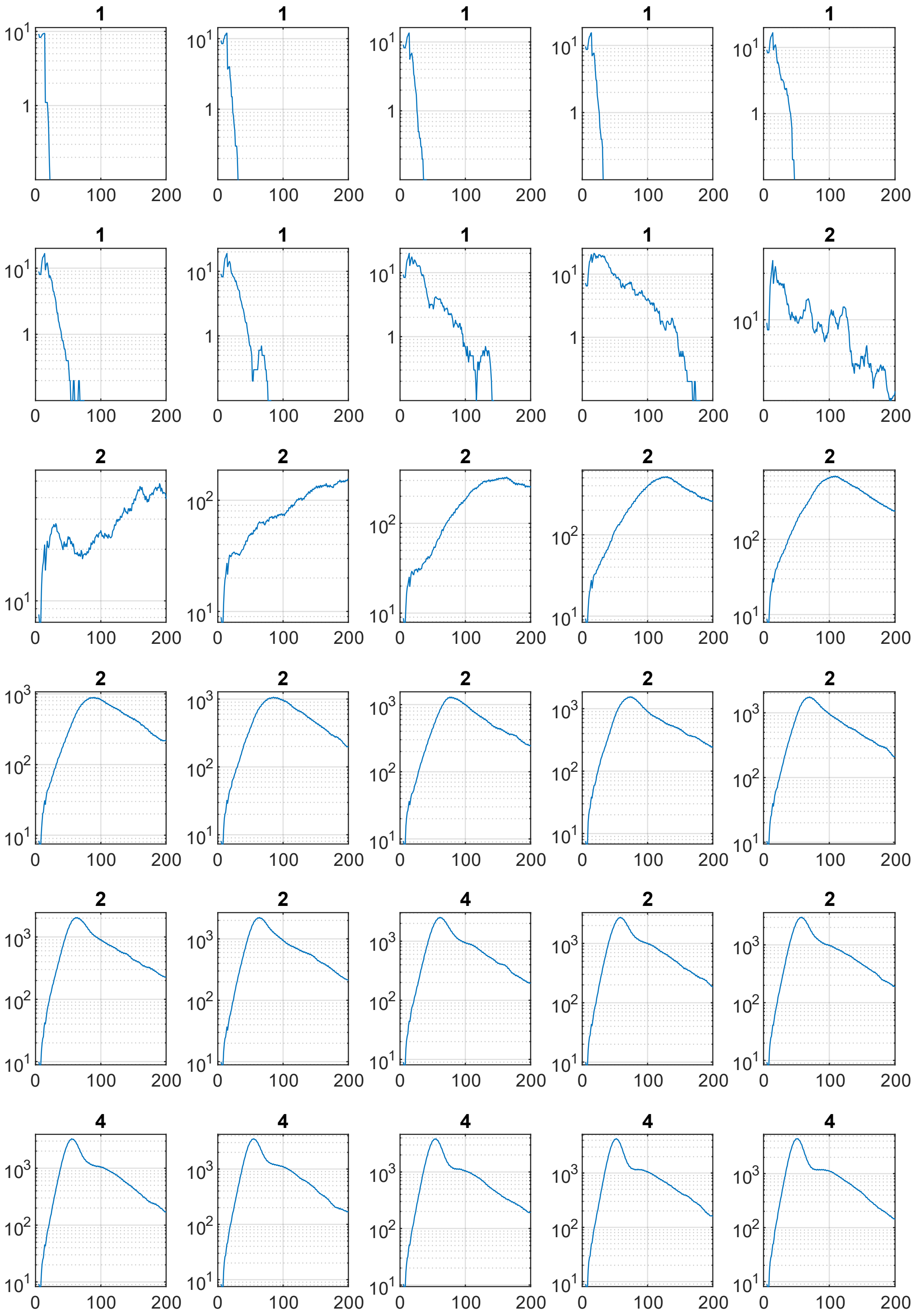


...

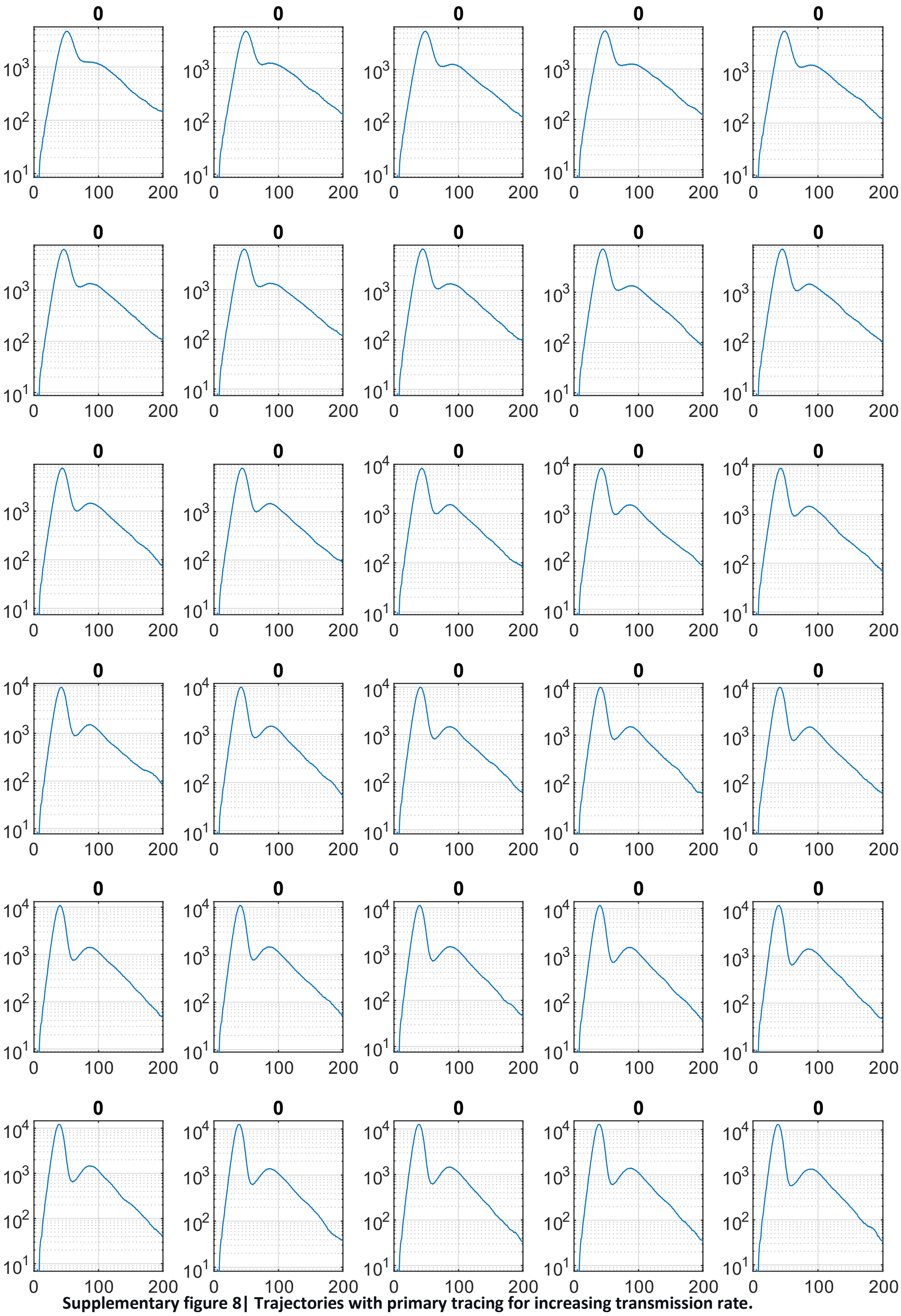

**Supplementary figure 8| Trajectories with primary tracing for increasing transmission rate.** Each Panel is a trajectory. The intercommunity links for all panels is 10%. The transmission rate, r, is [0.001:0.002:0.12] from left to right. The trajectories are labeled as 0 for “growth”, 1 for eradication, 2 for stability, 3 for oscillatory stability, 4 for stable after a large wave.

Supplementary Figure 9

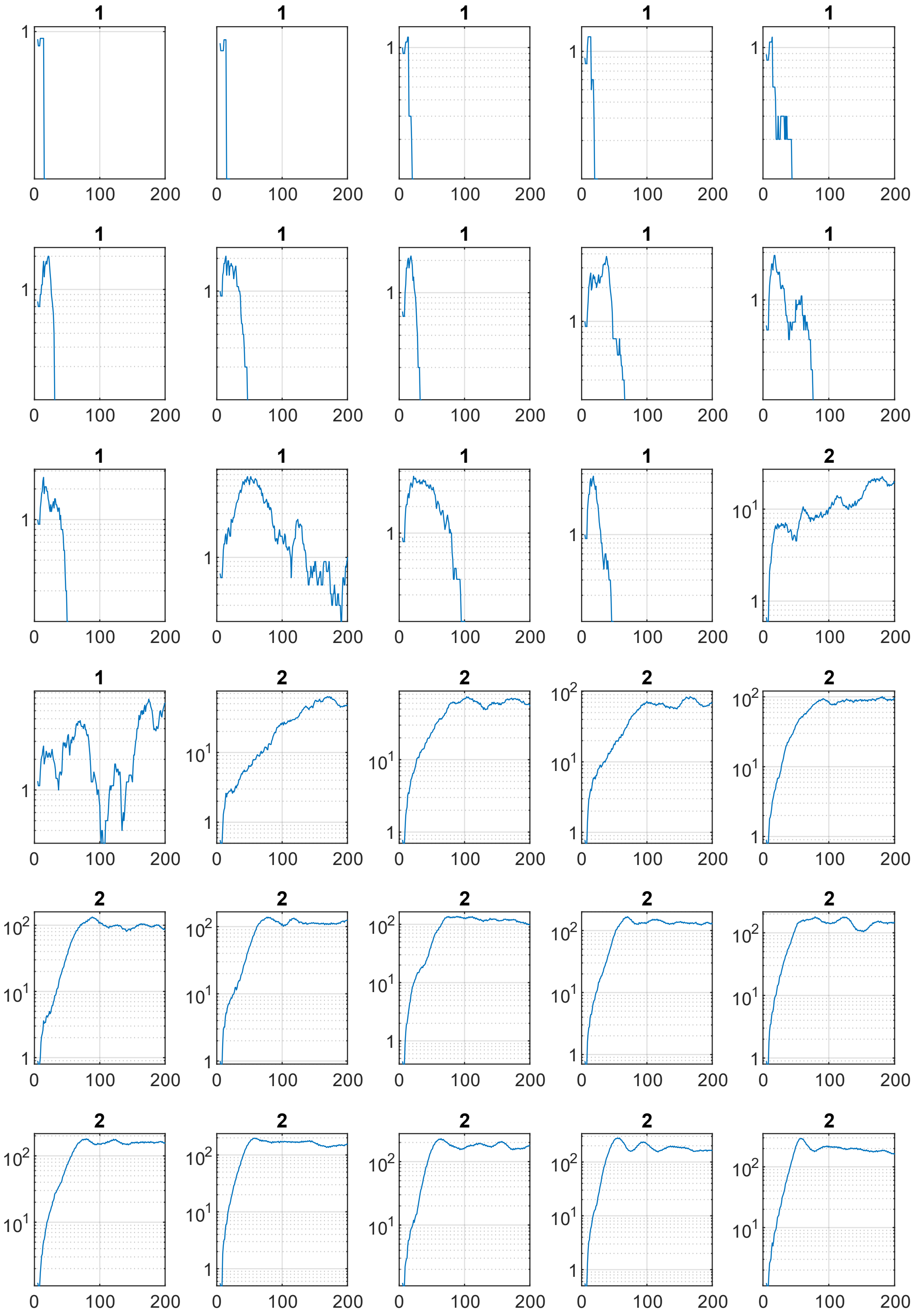

…

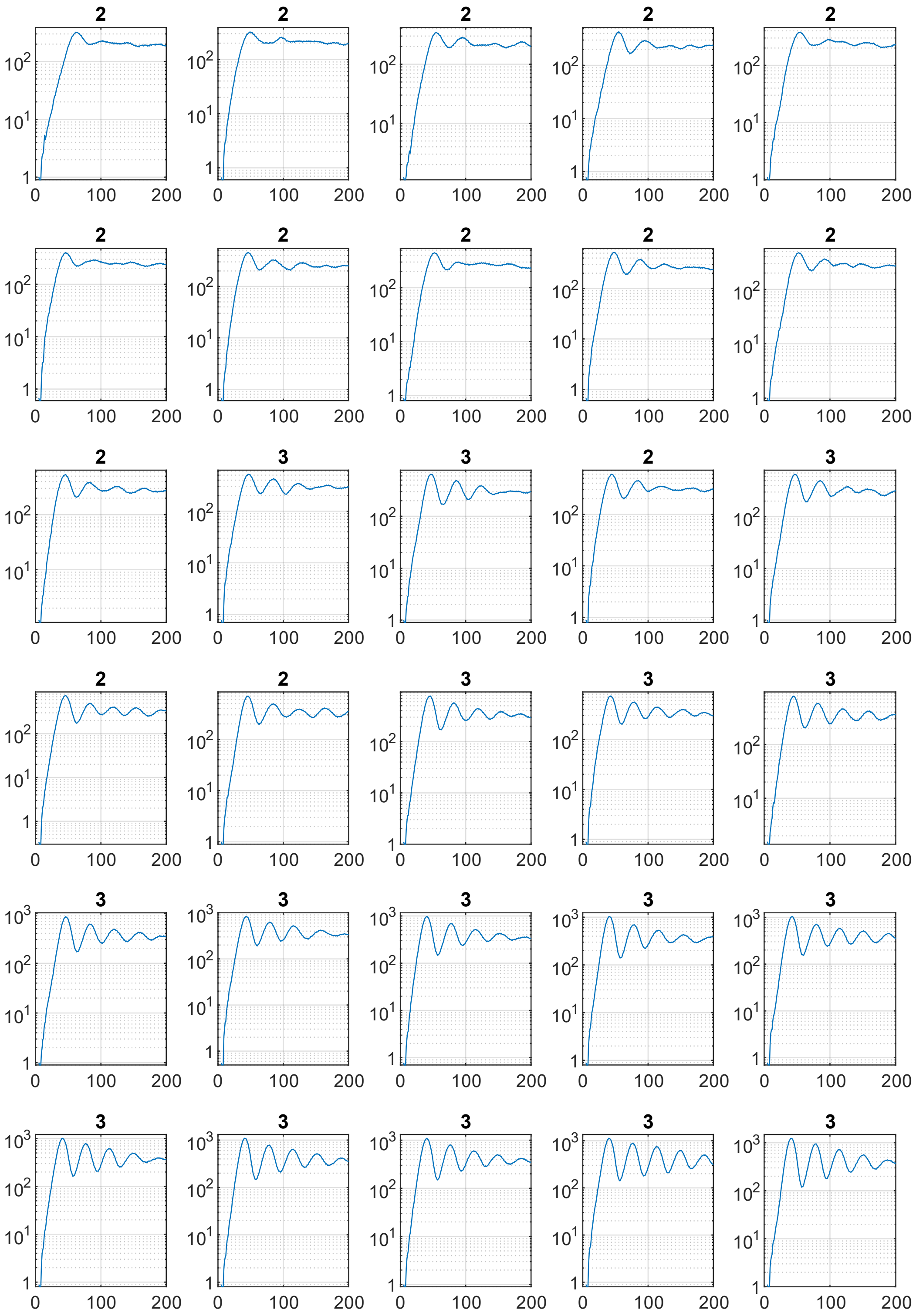


**Supplementary figure 9| Trajectories with secondary tracing for increasing transmission rate.** Each Panel is a trajectory. The intercommunity links for all panels is 10%. The transmission rate, r, is [0.001:0.002:0.12] from left to right. The trajectories are labeled as 0 for “growth”, 1 for eradication, 2 for stability, 3 for oscillatory stability, 4 for stable after a large wave.

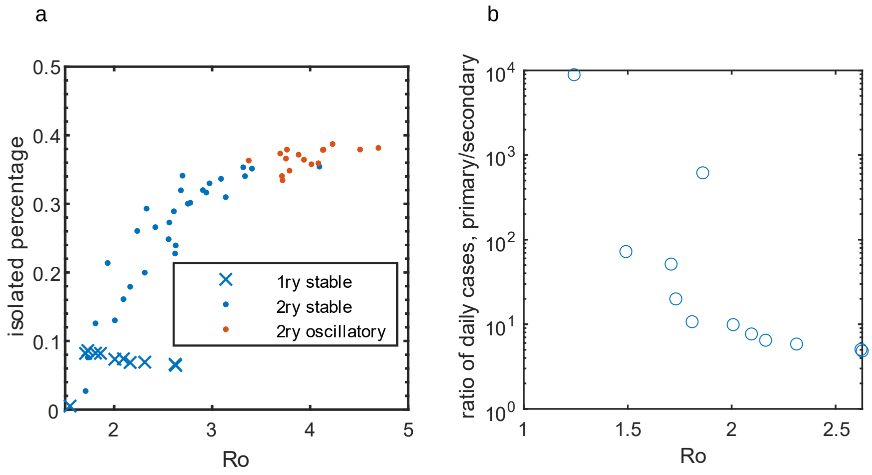


**Supplementary figure 10| Isolation percentages and the effect of tracing radius. a**, Isolated percentage of the population for primary and secondary tracing. The secondary tracing is significantly higher than the primary tracing. **b,** Number of daily cases for stable trajectories were estimated. The values from the primary tracing trajectories are divided by the values from the trajectories with secondary tracing. The range varies greatly. The ratio approaches around 6 for higher Ro.

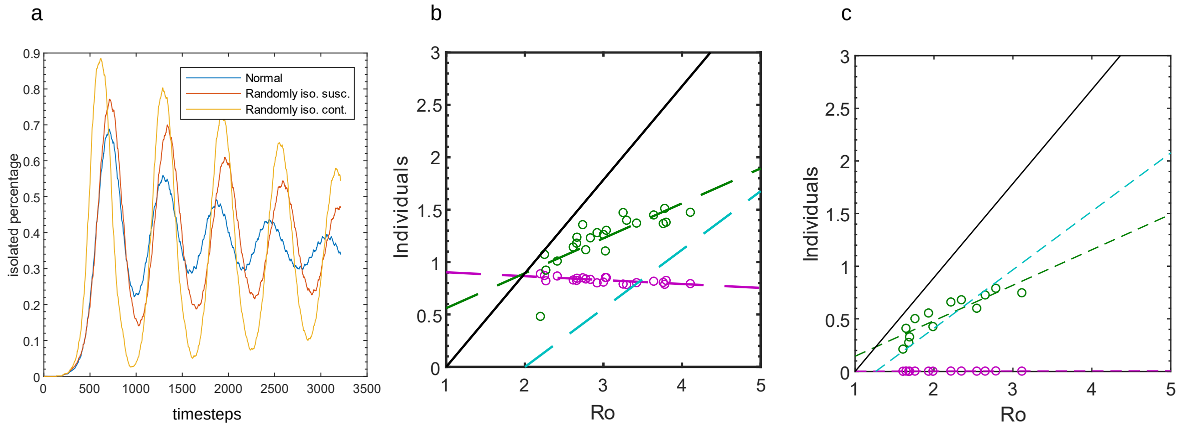


**Supplementary figure 11| Isolating susceptible contacts at random is a detriment to stability.** Two additional cases were simulated: 1) instead of isolating susceptible contacts, they are isolated at random, 2) all contacts are chosen at random **a**, Isolated percentage of the population. Randomly chosen trajectories shows higher oscillations. b, Successful isolations of non-descendants and the descendants are the same as the normal trajectories, Fig. 2f. Randomly choosing susceptibles can still impact the dynamics. Therefore, even the susceptibles of contacts are different from other susceptibles. Perhaps they have a higher probability of being infected. **c**, Isolating non-descendants and descendants are greatly impacted when all contacts are chosen at random.

Supplementary Figure 12

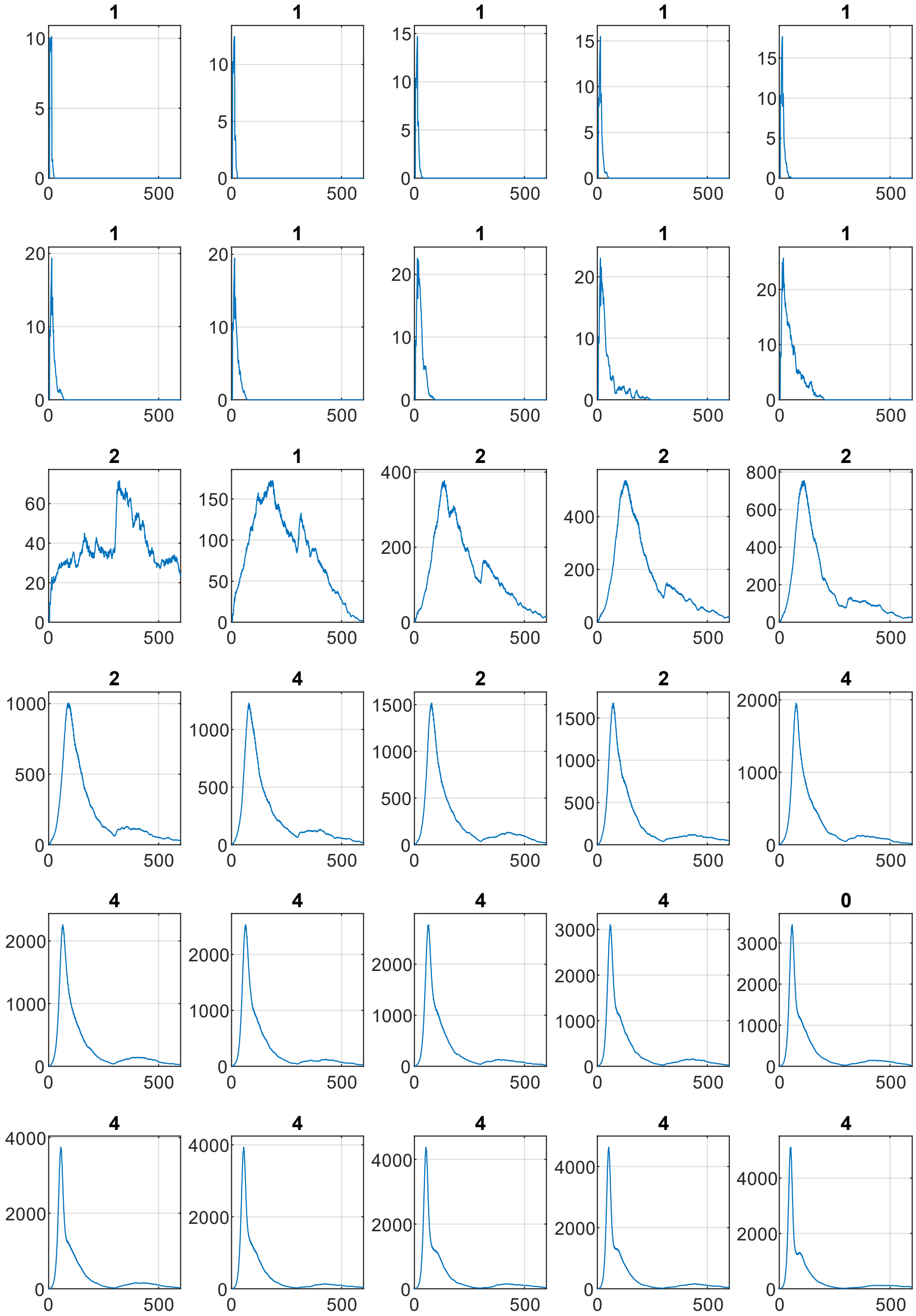


...

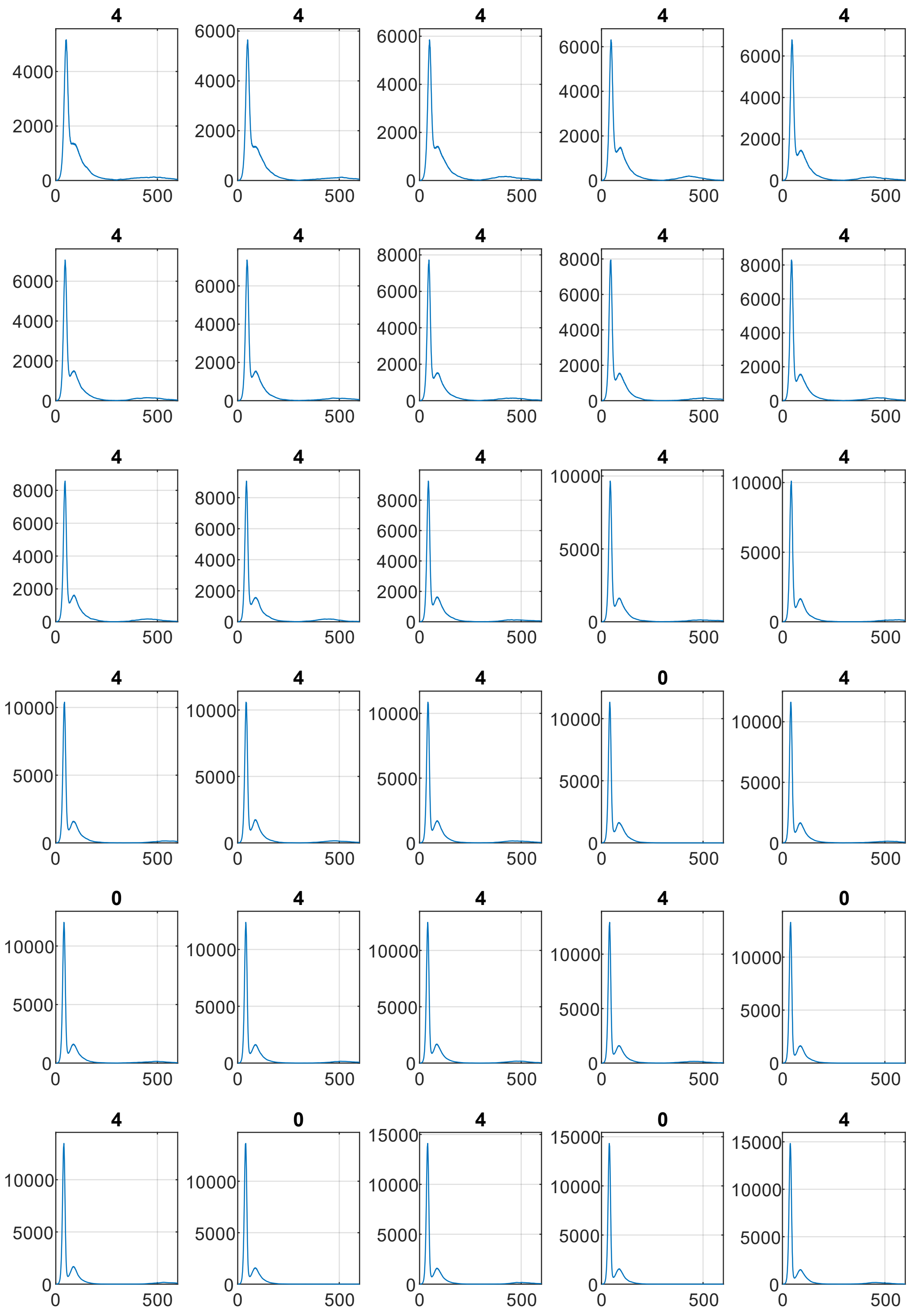


**Supplementary figure 12| Effect of mobility on trajectories with primary tracing.** Each Panel is a trajectory. The intercommunity links for all panels is 10%. The transmission rate, r, is [0.001:0.002:0.12] from left to right. The trajectories are labelled as 0 for “growth”, 1 for eradication, 2 for stability, 3 for oscillatory stability, 4 for the stable after a large wave. At 200 days, the population is shuffled for a day.

Supplementary Figure 13

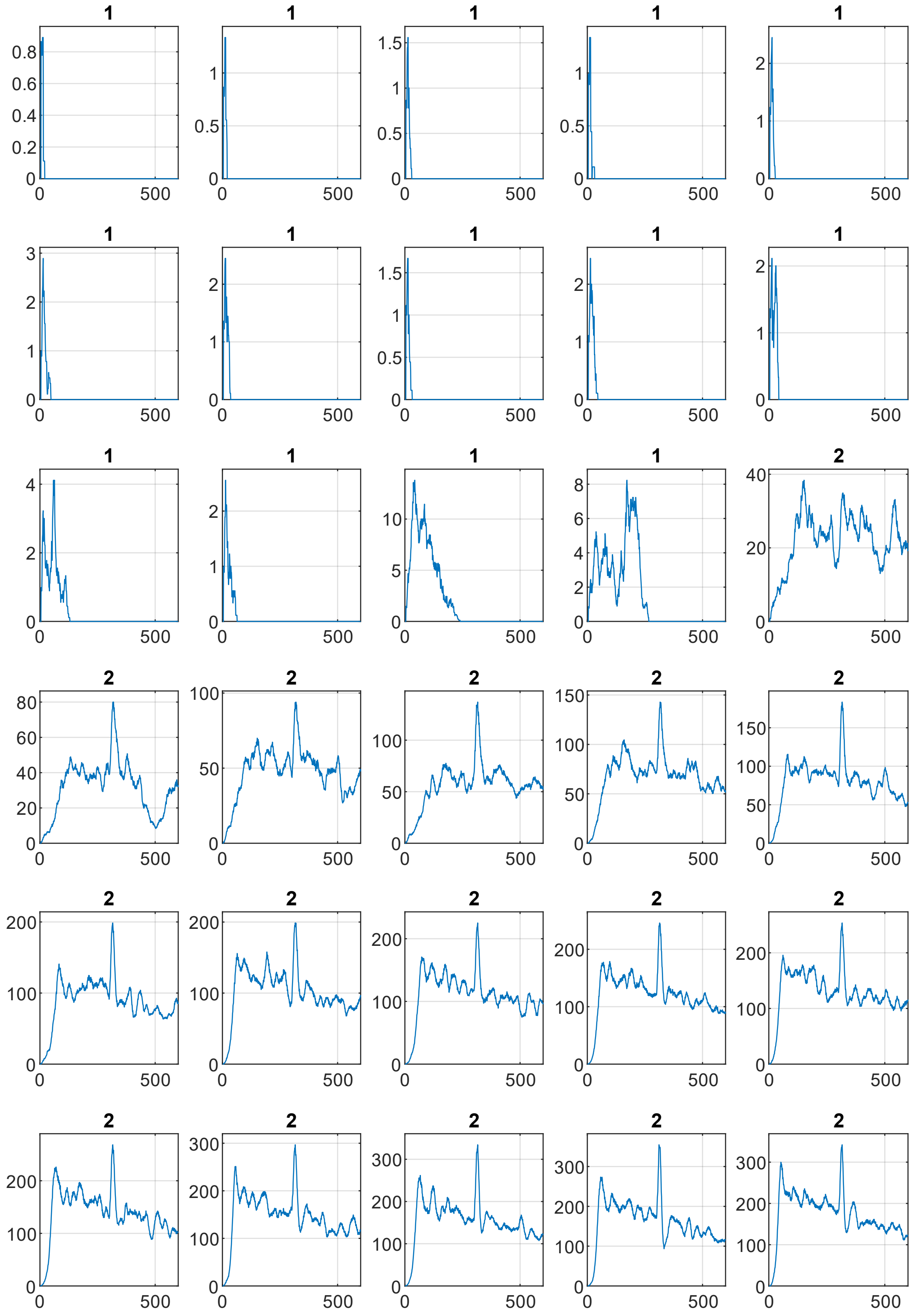


…

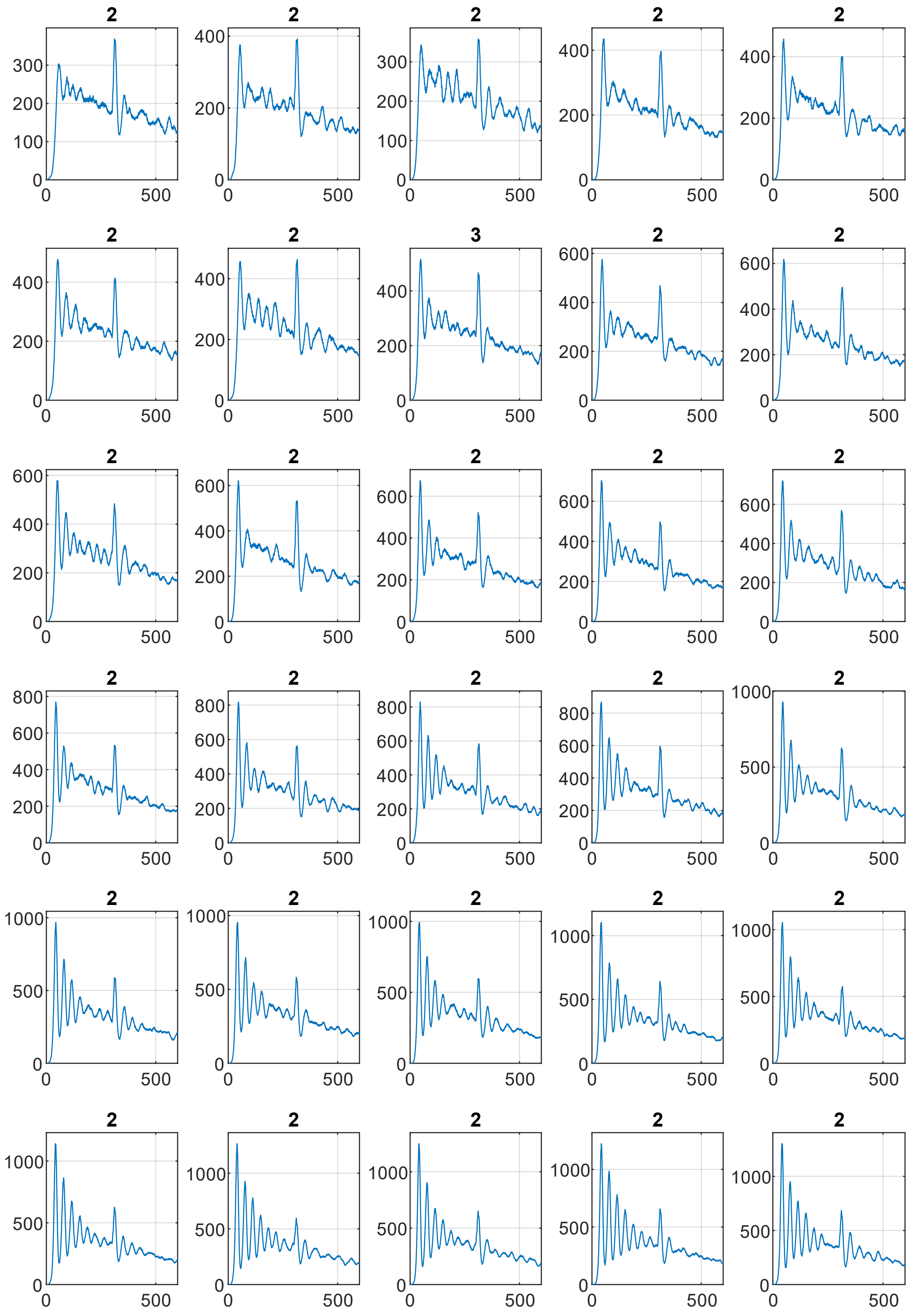


**Supplementary figure 13| Effect of mobility on trajectories with secondary tracing.** Each Panel is a trajectory. The intercommunity links for all panels is 10%. The transmission rate, r, is [0.001:0.002:0.12] from left to right. The trajectories are labeled as 0 for “growth”, 1 for eradication, 2 for stability, 3 for oscillatory stability, 4 for stable after a large wave. At 200 days, the population is shuffled for a day.

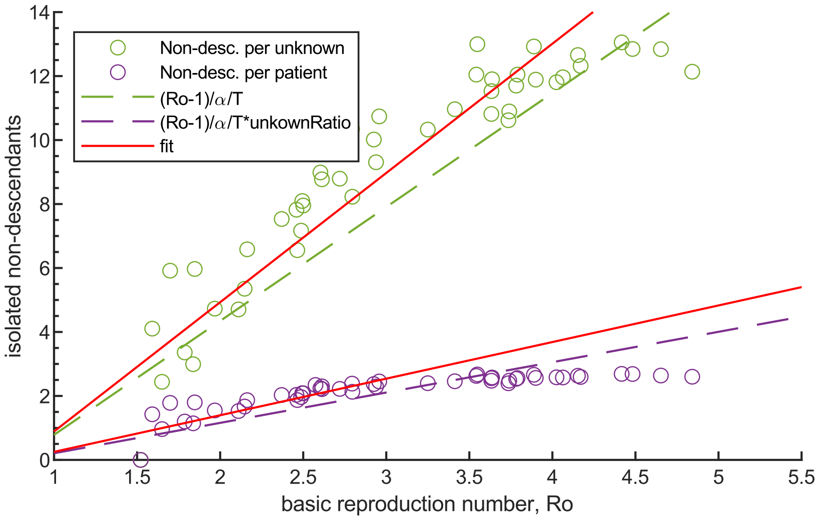


**Supplementary figure 14| Numerical assessment of required isolation for stability.** To determine the required number of non-descendant isolations, I simulated a case where susceptible contacts are not isolated. Thus, only non-descendant isolation matters. The values are directly observed from the trajectories with increasing Ro. The Ro is the basic reproduction number without any contact-tracing. In this agent-based model, the isolated individuals' contacts are also traced if they develop symptoms. This seemingly unimportant decision reduces the isolation of non-descendants for stability by 3 folds per documented patient. The documented patients from the isolated pool can still provide removal of non-descendants.

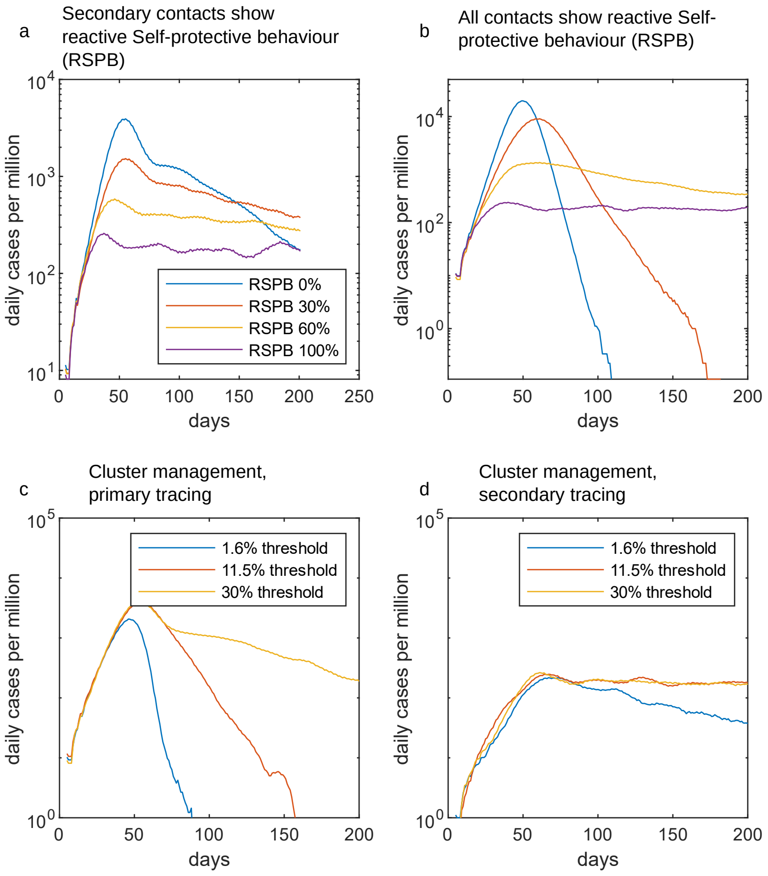


**Supplementary figure 15| Effect of reactive self-protection and cluster management on stability.** Reactive self-protection is modelled as a transmission rate reduction. **a**, Only secondary contacts get transmission penalty. The primary contacts are dealt with as before. **b**, All contacts get transmission penalty. Contacts are not isolated at all. **c**, Cluster management for simulations with primary tracing. **d**, Cluster management for simulations with secondary tracing.

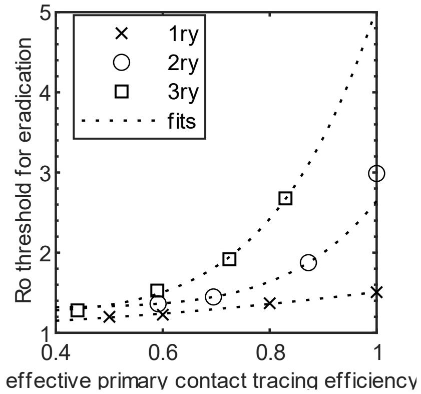


**Supplementary figure 15| Eradication thresholds are similar for modest effective primary contact tracing.** High order tracing increases the tracing success of the primary contacts. Eradication threshold is comparable at intermediate values.
